# Effectiveness of the 2023-2024 Omicron XBB.1.5-containing mRNA COVID-19 vaccine (mRNA-1273.815) in preventing COVID-19-related hospitalizations and medical encounters among adults in the United States: An interim analysis

**DOI:** 10.1101/2024.04.10.24305549

**Authors:** Hagit Kopel, Andre B. Araujo, Alina Bogdanov, Ni Zeng, Isabelle Winer, Jessamine Winer-Jones, Tianyi Lu, Morgan A. Marks, Mac Bonafede, Van Hung Nguyen, David Martin, James A. Mansi

**Affiliations:** Moderna, Inc., Cambridge, MA; Veradigm, Chicago, IL, USA; VHN Consulting Inc., Montreal, Canada

**Keywords:** mRNA-1273.815, Omicron XBB.1.5 vaccine, COVID-19 vaccine, vaccine effectiveness, COVID-19, hospitalizations, 2023-2024 COVID-19 vaccine

## Abstract

**Background/Objectives:** COVID-19 continues to pose a significant burden that impacts public health and the healthcare system as the SARS-CoV-2 virus continues to evolve. Regularly updated vaccines are anticipated to boost waning immunity and provide protection against circulating variants. This study evaluated vaccine effectiveness (VE) of mRNA-1273.815, a 2023-2024 Omicron XBB.1.5-containing mRNA COVID-19 vaccine, at preventing COVID-19-related hospitalizations and any medically attended COVID-19 in adults ≥18 years, overall, and by age and underlying medical conditions.

**Methods:** This retrospective cohort study used the Veradigm Network EHR linked to claims data to identify US adults ≥18 years of age who received the mRNA-1273.815 vaccine (exposed) matched 1:1 to individuals who did not receive a 2023-2024 updated COVID-19 vaccine (unexposed). Patients in the unexposed cohort were randomly matched to eligible mRNA-1273.815 recipients. Inverse probability of treatment weighting was used to adjust for differences between the two cohorts. The exposed cohort was vaccinated between September 12, 2023, and December 15, 2023, and individuals in both cohorts were followed up for COVID-19-related hospitalizations and medically attended COVID-19 until December 31, 2023. A Cox regression model was used to estimate the hazard ratio (HR). VE of the mRNA-1273.815 vaccine in preventing COVID-19-related hospitalizations and any medically attended COVID-19 was estimated as 100*(1-HR). Subgroup analyses were performed for adults ≥50, adults ≥65, and individuals with underlying medical conditions associated with severe COVID-19 outcomes.

**Results:** Overall, 859,335 matched pairs of mRNA-1273.815 recipients and unexposed adults were identified. The mean age was 63 years, and 80% of the study population was ≥50 years old. 61.5% of the mRNA-1273.815 cohort and 66.4% of the unexposed cohort had an underlying medical condition. Among the overall adult population (≥18 years), VE was 60.2% (53.4–66.0%) against COVID-19-related hospitalization and 33.1% (30.2%–35.9%) against medically attended COVID-19 over a median follow-up of 63 (IQR: 44–78) days. VE estimates by age and underlying medical conditions were similar.

**Conclusions:** These results demonstrate the significant protection provided by mRNA-1273.815 against COVID-19-related hospitalizations and any medically attended COVID-19 in adults 18 years and older, regardless of their vaccination history, and support CDC recommendations for vaccination with the 2023-2024 Omicron XBB.1.5-containing COVID-19 vaccine to prevent COVID-19-related outcomes, including hospitalizations.

## INTRODUCTION

COVID-19 continues to be the most common cause of severe outcomes due to viral respiratory illness in the United States (US) and was associated with over 900,000 hospitalizations and 75,000 deaths in 2023.^1–3^ In 2023, the weekly incidence of COVID-19-associated hospitalizations and deaths surpassed the combined weekly incidence due to influenza or respiratory syncytial virus (RSV), according to data from the Centers for Disease Control and Prevention (CDC).^1^ The burden of COVID-19 is not limited to the acute phase, as even patients with mild disease can experience symptoms and develop conditions that can last months following infection, including long COVID, and might require comprehensive care. ^4–6^

Due to the emergence of novel SARS-CoV-2 viral variants and the observed impact on vaccine performance, the US Food and Drug Administration (FDA) approved and authorized an updated formulation of COVID-19 mRNA vaccines on September 11, 2023, and an adjuvanted protein formulation on October 3, 2023, that were designed to target the circulating omicron variant XBB.1.5 of SARS-CoV-2.^7^ On September 12, 2023, the Advisory Committee on Immunization Practices (ACIP) recommended the 2023-2024 Omicron XBB.1.5-containing COVID-19 vaccines for all individuals 6 months of age and older.^7^

Early data suggest that SARS-CoV-2 XBB.1.5-containing vaccines elicit robust and diverse neutralizing antibody responses against XBB.1.5 as well as other recent SARS-CoV-2 variants, including JN.1.^10,11^ In addition, a few studies have examined the vaccine effectiveness (VE) of the 2023-2024 Omicron XBB.1.5-containing mRNA COVID-19 vaccine.^12–16^ However, there is no existing data on the real-world effectiveness of the mRNA-1273.815 vaccine specifically. Assessment of VE is essential not only for regulatory agencies and vaccine policy makers but also for healthcare providers to provide recommendations and increase vaccine confidence among their patients. This study aimed to evaluate the VE of the mRNA-1273.815 vaccine against COVID-19-related hospitalizations and any medically attended COVID-19 in adults 18 years or older.

## METHODS

### Data Sources

This study leveraged electronic health record (EHR) data from the Veradigm Network EHR linked to healthcare claims sourced from Komodo Health spanning March 1, 2020, through December 31, 2023. The EHR dataset consists of de-identified patient records sourced from ambulatory/outpatient primary care and specialty settings. The insurance claims data contains de-identified inpatient, outpatient, and pharmacy claims. This integrated claims and EHR dataset have been previously characterized by Boikos et al. and used previously in COVID-19 epidemiology and VE research.^2,17–19^

Patient records from multiple contributing data sources were linked using patient-level de-identified tokens, which were created deterministically from fields like name, date of birth, and sex using an algorithm developed by Datavant. The final linked dataset was created as a merge of the patient-level de-identified tokens in each individual dataset and contains no personal health information. Veradigm research staff were not involved in the preparation of datasets containing protected health information (PHI) or the actual running of the linkage algorithm.

The linked dataset used in this study contains only de-identified data as per the de-identification standard defined in Section §164.514(a) of the Health Insurance Portability and Accountability Act of 1996 (HIPAA) Privacy Rule. As a noninterventional, retrospective database study using data from a certified HIPAA– compliant de-identified research database, approval by an institutional review board was not required.

### Study Design Overview

We conducted a retrospective, non-interventional cohort study of mRNA-1273.815 VE in adults who were ≥18 years old and residing in the United States (US). Adults who received the mRNA-1273.815 vaccine during the vaccine intake period were identified and matched in a 1:1 ratio to adults with no evidence of receiving a 2023-2024 Omicron XBB.1.5-containing COVID-19 vaccine. Baseline patient characteristics (covariates) were measured, and inverse probability of treatment weighting (IPTW) was used to further reduce potential confounding by creating a weighted study sample based on the pre-defined covariates. Individuals in the vaccinated (exposed) and unvaccinated (unexposed) cohorts were followed for the two key outcomes, COVID-19-related hospitalizations and any medically attended COVID-19, during the follow-up period. Cox regression models were used to calculate hazard ratios (HR) and 95% confidence intervals (95% CI), which were converted to VE estimates. The study design is shown in Figure 1.

**Figure 1.**
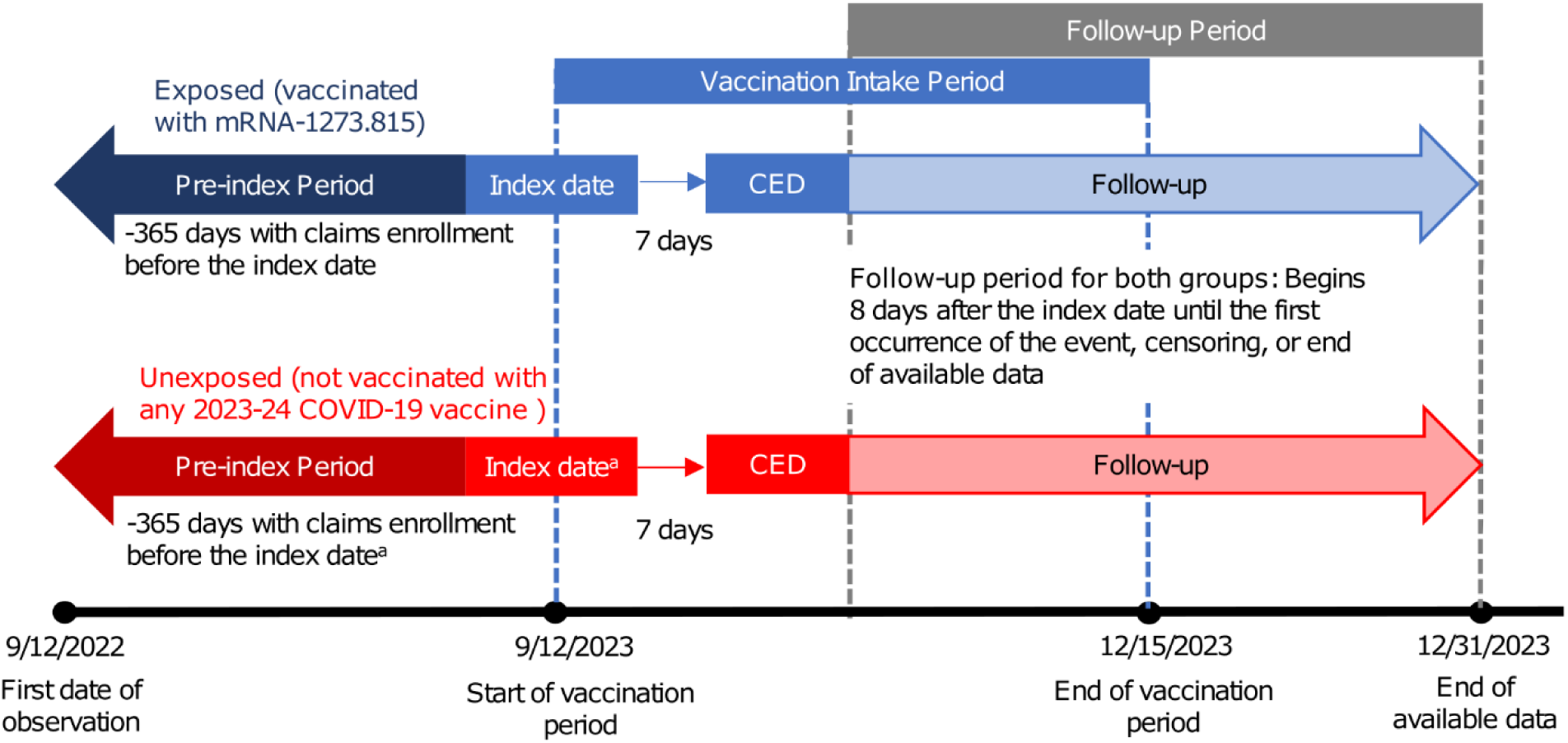
Study Design. CED, cohort entry date. ^a^The index date for unexposed individuals was assigned based on their 1:1 match in the exposed cohort.

### Study Population

Individuals with a record of vaccination with mRNA-1273.815 (50 mcg) during the vaccine intake period (September 12, 2023–December 15, 2023) were eligible for inclusion in the exposed cohort. Individuals without a record of vaccination with any 2023-2024 updated COVID-19 vaccine during the vaccine intake period were eligible for inclusion in the unexposed cohort.

The index date of the individuals in the exposed cohort was the date of their first mRNA-1273.815 vaccination. Individuals in the unexposed cohort were assigned multiple potential index dates based on the index dates of all possible direct matches in the exposed cohort using the following matching criteria: age on September 12, 2023 (exact match), sex (exact match), race (exact match), ethnicity (exact match), region (exact match), week of last claims or EHR activity, and receipt of a bivalent BA.4/BA.5 COVID-19 vaccine between Sept 1, 2022, and September 11, 2023 (yes/no). Unexposed individuals with no potential matches in the exposed cohort were excluded.

We required that individuals have continuous enrollment in medical and pharmacy claims from September 12, 2022, through 7 days after the index date (cohort entry date [CED]). Individuals with any of the following were excluded: 1) missing sex or region data, 2) evidence of COVID-19 diagnosis or treatment between 90 days before and 7 days after the index date, or 3) evidence of vaccination with any COVID-19 vaccine in the 60 days prior to the index day through 7 days after the index date (excluding the index vaccination in the exposed group). Age was a required field in the underlying data file. In addition, we required that individuals have at least 1 day of follow-up after the CED. Follow-up began the day after the CED and continued until the earliest of the following: outcome of interest, the end of continuous enrollment, receipt of a subsequent COVID-19 vaccine, or end of the follow-up period (December 31, 2023).

After the above selection criteria were applied, the final matches were created as follows. First, an individual in the exposed cohort was randomly selected. Then, if they had exactly one match in the unexposed cohort who met all the selection criteria, they were assigned that individual as a match, and both were removed from further matching. If they had multiple potential matches who met all the selection criteria, they were randomly assigned one match, and both were removed from further matching. If the exposed individual had zero remaining matches who met all the selection criteria, they were excluded from the study. This process was repeated until all individuals in the exposed cohort were either matched to someone in the unexposed cohort or excluded from the study. Individuals in the unexposed cohort who were not matched to an individual in the exposed cohort were excluded. The final index date for individuals in the unexposed cohort was the same as their matched mRNA-1273.815 counterpart.

The final study population included only adults ≥18 years old who met the eligibility criteria. The following subsets of the overall study population were defined for exploratory analyses: adults ≥50 years old, adults ≥65 years old, and adults ≥18 years old with at least one underlying medical condition that increased the risk of severe outcomes from COVID-19 as defined by the CDC (see also the covariates subsection).^20^ The codes used to identify 2023-2024 Omicron XBB.1.5-containing COVID-19 vaccines, including the mRNA-1273.815 vaccine, are reported in Supplementary Table 1. The codes used to identify COVID-19 vaccination are reported in Supplementary Table 2. The codes used to identify COVID-19 diagnosis or treatment are reported in Supplementary Table 3.

### Outcome Measures

The primary outcome of interest was COVID-19-related hospitalization, and the secondary outcome of interest was any medically attended COVID-19, which included various clinical settings such as emergency department visits, urgent care visits, office visits, telemedicine visits, laboratory results, etc. Each outcome was evaluated independently so that an individual could be counted as having both a COVID-19-related hospitalization and medically attended COVID-19.

COVID-19-related hospitalizations were identified from hospitalization claims with documentation of COVID-19 diagnosis in any position. Any medically attended COVID-19 were identified from any EHR or medical claim in any position with either a COVID-19 diagnosis or positive laboratory test result. Laboratory test results came only from the EHR.

The codes used to identify COVID-19 diagnosis are reported in Supplementary Table 3. Only diagnosis codes were used to identify COVID-19-related hospitalizations. Both diagnosis and positive lab-test results were used to identify any medically attended COVID-19.

### Covariates

The study utilized covariate information including demographics (age on September 12, 2023, sex (Female, Male), race (Black, White, other, and unknown), ethnicity (Hispanic, non-Hispanic, and unknown)), insurance type (Commercial, Medicaid, Medicare Advantage, Medicare Fee-For-Service, other, and unknown), region (Midwest, Northeast, South, West, and unknown), and month of index. History of COVID-19 illness and vaccination were captured as well. The time since last COVID-19 vaccination and time since last COVID-19 infection were captured using a look-back period that started on March 1, 2020. The number of outpatient visits and the number of hospitalizations were captured using a 12-month pre-period. In addition, the presence of underlying medical conditions associated with an increased risk of severe COVID-19 outcomes, as defined by the US CDC, was captured.^20^ These included asthma, cancer, cerebrovascular disease, chronic kidney disease, chronic liver disease, chronic lung disease, cystic fibrosis, dementia, diabetes mellitus, disability, heart conditions, human immunodeficiency virus (HIV), mental health conditions, obesity (body mass index > 30), physical inactivity, pregnancy, primary immunodeficiencies, respiratory tuberculosis, smoking, and solid organ or stem cell transplant, and use of select immunosuppressive medications. With the exception of pregnancy, underlying medical conditions were captured in the 12-month pre-index period. Pregnancy was captured in the 301 days preceding the index date. Code sets and medication lists used to identify underlying medical conditions are listed in Supplementary Table 4. Patients with at least one of these medical conditions were considered “high-risk” for severe COVID-19 outcomes.

### Data Analysis

Propensity scores predicting receipt of mRNA-1273.815 were calculated for each subject using a multivariable logit model adjusted for all covariates outlined above. Stabilized and truncated weights were used to re-weigh the study sample using the IPTW methodology. Sample balance before and after IPTW was assessed by calculation of the standardized mean differences (SMD). SMDs with absolute values greater than 0.1 indicated covariate imbalance. Covariates were reported descriptively before and after weighting with means and standard deviations (SD) for continuous variables and N and percent for categorical variables.

Unadjusted and adjusted HR were calculated for each outcome. Unadjusted HRs were calculated using a Cox regression model with mRNA-1273.815 vaccination as the only predictor. Adjusted HRs were calculated using a weighted Cox regression model that includes mRNA-1273.815 vaccination and any covariates with an SMD greater than 0.1 after IPTW. Subsequently, the unadjusted and adjusted VE for each outcome were calculated as 100*(1-HR) and reported with 95% confidence intervals (Cis).

This analysis was conducted using SAS V9.4.

## RESULTS

Overall, this study included 1,718,670 individuals; 859,335 adults who received the mRNA-1273.815 vaccine between September 12, 2023, and December 15, 2023 (exposed cohort), met all the study criteria and were directly matched 1:1 to an unexposed adult (Figure 2). Of these, 70.5% (N = 605,816) of each cohort had previously received the COVID-19 bivalent vaccine. The exposed and unexposed cohorts were well balanced after weighting, with no SMDs greater than 0.1. Therefore, there was no need for additional adjustment for covariates in the weighted Cox regression model. After weighting, the mean (SD) age in both cohorts was 63 (16) years; 57% were female, 60% were White, 6% were Black, and more than 80% received their most recent COVID-19 vaccine more than 180 before the index date (Table 1). Approximately 80% (N = 686,135) of the study population was at least 50 years old, and 54% (N = 465,061) were at least 65 years old (Table 2).

**Figure 2.**
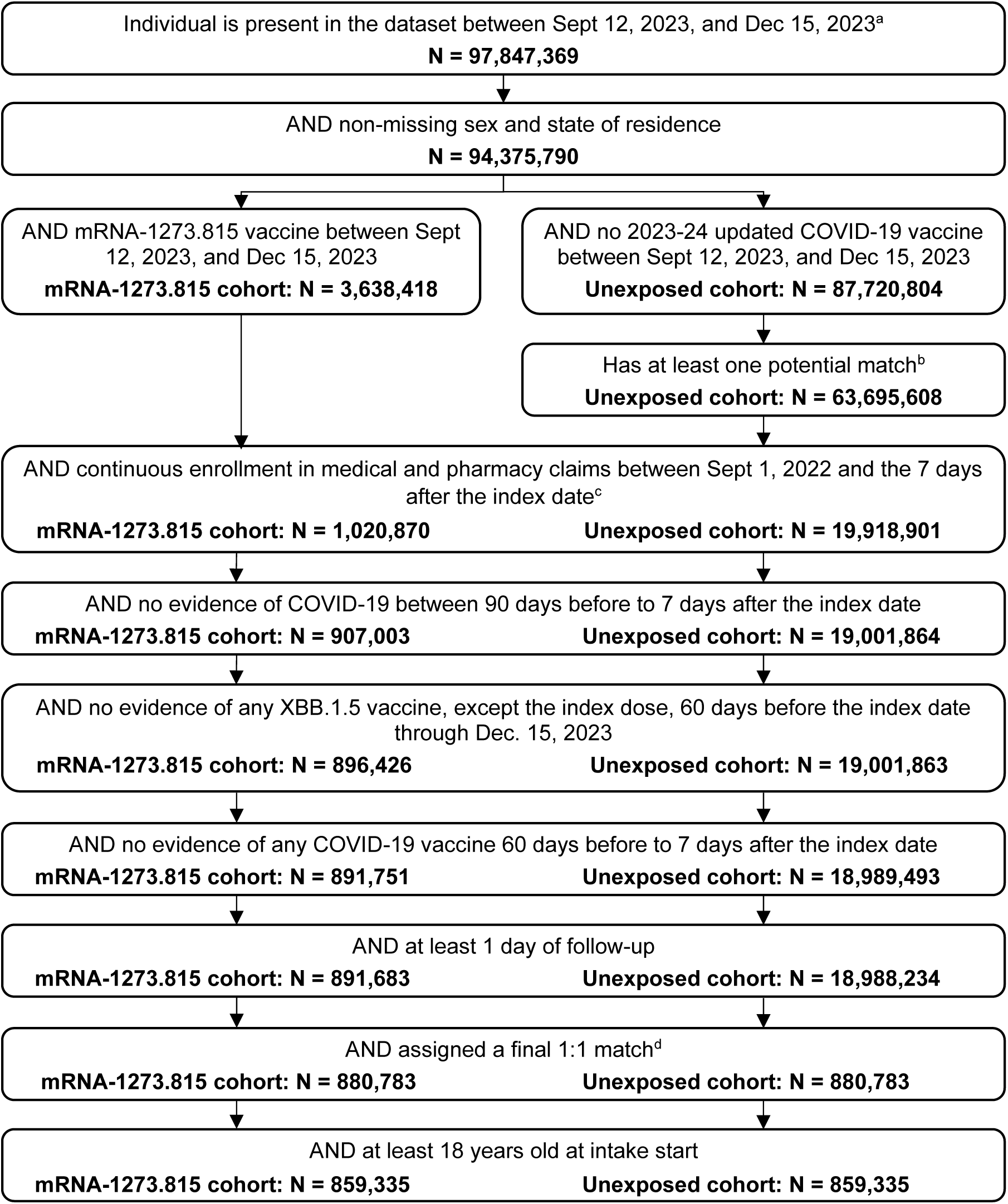
Patient Selection. EHR, electronic health records ^a^Includes individuals with continuous enrollment for any portion of this period along with those who have at least one record in claims or EHR during this period. ^b^Matched on age on Sept 12, 2023, sex, race, ethnicity, region, latest activity week, and receipt of a bivalent COVID-19 vaccine between Sept 1, 2022, and Sept 11, 2023. ^c^For the mRNA-1273.815 cohort, the index date is the date of first evidence of mRNA-1273.815 vaccine. For the unexposed cohort, each patient was assigned potential index dates for evaluation based on their potential matches in the mRNA-1273.815 vaccine cohort in Line 4. Their final index date was assigned based on their match in Line 10. ^D^Among all potential unexposed cohort matches who made it through the waterfall, each individual in the mRNA-1273.815 cohort was randomly assigned exactly one final match. Individuals in the mRNA-1273.815 cohort who no longer had any potential matches in the unexposed cohort were excluded. Unmatched unexposed individuals were excluded.

**Table 1.** Baseline characteristics of individuals included as covariates in the main analysis of the primary and secondary objectives. Data are presented as n (%) unless otherwise stated.

|  |  | Pre-weighting |  |  | Post-weighting |  |  |
| --- | --- | --- | --- | --- | --- | --- | --- |
|  |  | mRNA-1273.815 | unexposed | SMD | mRNA-1273.815 | unexposed | SMD |
| Number of patients |  | 859,335 | 859,335 |  | 855,567 | 858,518 |  |
| Age at intake, mean (SD) |  | 63 (16.3) | 63 (16.3) | 0 | 63 (16.2) | 63 (16.3) | 0.003 |
| Sex, N (%) | Female | 490,534 (57.1%) | 490,534 (57.1%) | 0 | 487,795 (57.0%) | 489,828 (57.1%) | <.001 |
|  | Male | 368,801 (42.9%) | 368,801 (42.9%) |  | 367,772 (43.0%) | 368,690 (42.9%) |  |
| Race, N (%) | Black | 50,444 (5.9%) | 50,444 (5.9%) | 0 | 50,431 (5.9%) | 50,585 (5.9%) | 0.002 |
|  | Other | 129,943 (15.1%) | 129,943 (15.1%) |  | 128,936 (15.1%) | 129,625 (15.1%) |  |
|  | White | 519,496 (60.5%) | 519,496 (60.5%) |  | 518,019 (60.5%) | 519,255 (60.5%) |  |
|  | Unknown | 159,452 (18.6%) | 159,452 (18.6%) |  | 158,181 (18.5%) | 159,054 (18.5%) |  |
| Ethnicity, N (%) | Hispanic | 19,228 (2.2%) | 19,228 (2.2%) | 0 | 19,161 (2.2%) | 19,230 (2.2%) | <.001 |
|  | Non-Hispanic | 506,284 (58.9%) | 506,284 (58.9%) |  | 504,390 (59.0%) | 506,042 (58.9%) |  |
|  | Unknown | 333,823 (38.8%) | 333,823 (38.8%) |  | 332,016 (38.8%) | 333,247 (38.8%) |  |
| Insurance Type <sup>a</sup>, N (%) | Commercial | 359,650 (41.9%) | 331,467 (38.6%) | 0.252 | 345,687 (40.4%) | 345,777 (40.3%) | 0.014 |
|  | Medicaid | 45,032 (5.2%) | 103,319 (12.0%) |  | 70,771 (8.3%) | 74,236 (8.6%) |  |
|  | Medicare Advantage | 346,959 (40.4%) | 316,761 (36.9%) |  | 331,133 (38.7%) | 330,519 (38.5%) |  |
|  | Medicare FFS | 7,486 (0.9%) | 12,455 (1.4%) |  | 9,825 (1.1%) | 9,960 (1.2%) |  |
|  | Other | 96,309 (11.2%) | 90,716 (10.6%) |  | 93,889 (11.0%) | 93,764 (10.9%) |  |
|  | Unknown | 3,899 (0.5%) | 4,617 (0.5%) |  | 4,262 (0.5%) | 4,263 (0.5%) |  |
| Region, N (%) | Midwest | 201,209 (23.4%) | 201,209 (23.4%) | 0 | 200,289 (23.4%) | 200,933 (23.4%) | <.001 |
|  | Northeast | 212,504 (24.7%) | 212,504 (24.7%) |  | 211,548 (24.7%) | 212,299 (24.7%) |  |
|  | South | 261,628 (30.4%) | 261,628 (30.4%) |  | 260,827 (30.5%) | 261,549 (30.5%) |  |
|  | West | 183,978 (21.4%) | 183,978 (21.4%) |  | 182,887 (21.4%) | 183,722 (21.4%) |  |
|  | Unknown | 16 (0.0%) | 16 (0.0%) |  | 16 (0.0%) | 16 (0.0%) |  |
|  |  | mRNA-1273.815 | unexposed | SMD | mRNA-1273.815 | unexposed | SMD |
| Month of index, N (%) | Sept 2023 | 131,504 (15.3%) | 131,504 (15.3%) |  | 127,961 (15.0%) | 132,666 (15.5%) |  |
|  | Oct 2023 | 435,505 (50.7%) | 435,505 (50.7%) |  | 431,090 (50.4%) | 435,894 (50.8%) |  |
|  | Nov 2023 | 230,482 (26.8%) | 230,482 (26.8%) |  | 233,467 (27.3%) | 228,676 (26.6%) |  |
|  | Dec 2023 | 61,844 (7.2%) | 61,844 (7.2%) |  | 63,050 (7.4%) | 61,283 (7.1%) |  |
| Number of outpatient visits, mean (SD) |  | 2.5 (6.1) | 2.8 (6.7) | 0.046 | 2.6 (6.5) | 2.6 (6.4) | <.001 |
| Number of hospitalizations, mean (SD) |  | 0.1 (0.5) | 0.2 (0.8) | **0.111** | 0.2 (0.7) | 0.2 (0.7) | 0.010 |
| Time since last COVID-19 vaccine, N (%) | 60- ≤90 days | 4,294 (0.5%) | 8,279 (1.0%) | **0.233** | 6,229 (0.7%) | 6,309 (0.7%) | 0.012 |
|  | 91–180 days | 48,897 (5.7%) | 33,964 (4.0%) |  | 41,057 (4.8%) | 40,557 (4.7%) |  |
|  | >180 days | 729,900 (84.9%) | 680,131 (79.1%) |  | 705,358 (82.4%) | 705,288 (82.2%) |  |
|  | Not reported | 76,244 (8.9%) | 136,961 (15.9%) |  | 102,924 (12.0%) | 106,364 (12.4%) |  |
| Time since last COVID-19 infection, N (%) | ≤120 days | 2,773 (0.3%) | 2,547 (0.3%) | 0.092 | 2,650 (0.3%) | 2,657 (0.3%) | 0.004 |
|  | 121–180 days | 4,659 (0.5%) | 4,453 (0.5%) |  | 4,529 (0.5%) | 4,550 (0.5%) |  |
|  | >180 days | 107,547 (12.5%) | 135,113 (15.7%) |  | 119,848 (14.0%) | 121,310 (14.1%) |  |
|  | Not reported | 744,356 (86.6%) | 717,222 (83.5%) |  | 728,540 (85.2%) | 730,002 (85.0%) |  |
| Patients with underlying medical conditions, N (%) | Asthma | 62,889 (7.3%) | 65,534 (7.6%) | 0.012 | 64,149 (7.5%) | 64,388 (7.5%) | <.001 |
|  | Cancer | 72,288 (8.4%) | 76,451 (8.9%) | 0.017 | 74,348 (8.7%) | 74,496 (8.7%) | <.001 |
|  | Cerebrovascular disease | 47,659 (5.5%) | 61,963 (7.2%) | 0.068 | 54,323 (6.3%) | 54,966 (6.4%) | 0.002 |
|  | Chronic kidney disease | 79,032 (9.2%) | 96,586 (11.2%) | 0.068 | 87,441 (10.2%) | 88,088 (10.3%) | 0.001 |
|  | Chronic lung disease | 65,155 (7.6%) | 85,061 (9.9%) | 0.082 | 74,755 (8.7%) | 75,485 (8.8%) | 0.002 |
|  | Chronic liver disease | 8,636 (1.0%) | 11,492 (1.3%) | 0.031 | 10,003 (1.2%) | 10,135 (1.2%) | 0.001 |
|  | Cystic fibrosis | 134 (0.0%) | 128 (0.0%) | <.001 | 128 (0.0%) | 129 (0.0%) | <.001 |
|  | Diabetes type 1 or 2 | 173,719 (20.2%) | 204,775 (23.8%) | 0.087 | 188,907 (22.1%) | 189,939 (22.1%) | 0.001 |
|  | Disability | 49,050 (5.7%) | 50,148 (5.8%) | 0.006 | 49,885 (5.8%) | 49,966 (5.8%) | <.001 |
|  | Heart conditions | 124,732 (14.5%) | 151,068 (17.6%) | 0.084 | 137,084 (16.0%) | 138,141 (16.1%) | 0.002 |
|  | HIV | 3,665 (0.4%) | 4,001 (0.5%) | 0.006 | 3,852 (0.5%) | 3,863 (0.4%) | <.001 |
|  | Mental health disorders | 123,072 (14.3%) | 142,194 (16.5%) | 0.062 | 132,719 (15.5%) | 133,482 (15.5%) | 0.001 |

|  | Pre-weighting |  |  | Post-weighting |  |  |
| --- | --- | --- | --- | --- | --- | --- |
|  | mRNA-1273.815 | unexposed | SMD | mRNA-1273.815 | unexposed | SMD |
| Neurological conditions | 20,390 (2.4%) | 33,239 (3.9%) | 0.086 | 26,394 (3.1%) | 26,993 (3.1%) | 0.003 |
| Obesity | 193,592 (22.5%) | 215,532 (25.1%) | 0.060 | 204,329 (23.9%) | 205,193 (23.9%) | <.001 |
| Primary immunodeficiencies | 10,061 (1.2%) | 11,991 (1.4%) | 0.020 | 11,051 (1.3%) | 11,077 (1.3%) | <.001 |
| Pregnancy | 1,189 (0.1%) | 1,316 (0.2%) | 0.004 | 1,203 (0.1%) | 1,239 (0.1%) | 0.001 |
| Physical inactivity | 616 (0.1%) | 856 (0.1%) | 0.010 | 734 (0.1%) | 743 (0.1%) | <.001 |
| Smoking | 96,971 (11.3%) | 122,476 (14.3%) | 0.089 | 109,043 (12.7%) | 110,116 (12.8%) | 0.002 |
| Solid organ or hematopoietic stem cell transplant | 2,583 (0.3%) | 2,934 (0.3%) | 0.007 | 2,802 (0.3%) | 2,796 (0.3%) | <.001 |
| Tuberculosis | 197 (0.0%) | 301 (0.0%) | 0.007 | 247 (0.0%) | 252 (0.0%) | <.001 |
| Use of immunosuppressants | 42,261 (4.9%) | 39,729 (4.6%) | 0.014 | 41,128 (4.8%) | 41,049 (4.8%) | 0.001 |

HIV, human immunodeficiency virus; SD, standard deviation; SMD, standardized mean difference
<sup>a</sup> Payers were evaluated in this order: Medicare FFS -> Medicare Advantage -> Medicaid -> Commercial ->Other -> Unknown. Individuals who were dual eligible for Medicare and Medicaid were counted as Medicare for the purpose of this study.

**Table 2.** Population and event counts during the variable follow-up period.

|  | All adults (≥18 years) |  | ≥18 years and high risk<sup>a</sup> |  | ≥50 years |  | ≥65 years |  |
| --- | --- | --- | --- | --- | --- | --- | --- | --- |
|  | mRNA-1273.815 | unexposed | mRNA-1273.815 | unexposed | mRNA-1273.815 | unexposed | mRNA-1273.815 | unexposed |
| Population counts, pre-IPTW | 859,335 | 859,335 | 528,465 | 570,783 | 686,135 | 686,135 | 465,061 | 465,061 |
| Population counts, post-IPTW | 855,567 | 858,518 | 526,294 | 570,226 | 683,370 | 685,233 | 463,172 | 464,267 |
| Follow-up time in days, median (IQR) | 63 (44–78) | 63 (44–78) | 64 (45–78) | 63 (44–77) | 64 (45–78) | 64 (45–78) | 65 (46–79) | 65 (46–79) |
| COVID-19-related hospitalization, counts (%), post-IPTW | 216 (0.03%) | 546 (0.06%) | 194 (0.04%) | 515 (0.09%) | 205 (0.03%) | 529 (0.08%) | 190 (0.04%) | 483 (0.10%) |
| Any medically-attended COVID-19, counts (%), post-IPTW | 3,448 (0.4%) | 5,181 (0.6%) | 2,621 (0.5%) | 4,367 (0.8%) | 2,974 (0.4%) | 4,613 (0.7%) | 2,184 (0.5%) | 3,581 (0.8%) |

IPTW, inverse probability of treatment weighting; IQR, interquartile range
<sup>a</sup> Defined by CDC: https://www.cdc.gov/coronavirus/2019-ncov/hcp/clinical-care/underlyingconditions.html

The majority of individuals, including 61.5% (N = 528,465) of the mRNA-1273.815 cohort and 66.4% (N = 570,783) of the unexposed cohort, had at least one underlying medical condition associated with an increased risk of severe COVID-19 outcomes (Table 2). The most common underlying medical conditions pre-weighting were obesity (exposed: 22.5% and unexposed: 25.1%), diabetes mellitus (20.2% and 23.8%, respectively), and heart conditions (14.5% and 17.6%, respectively) (Table 1). After matching on characteristics that included receipt of a bivalent COVID-19 vaccine, roughly 88% of each cohort had a record of previous COVID-19 vaccination.

The median follow-up time was 63 days for the overall analysis. The median follow-up time for the ≥50, ≥65, and high-risk cohorts were 64, 65, and 63 days, respectively (Table 2). For the overall population analysis, 762 COVID-19-related hospitalizations and 8,629 COVID-19-related medical encounters were identified in the post-weighing cohort. Of these, 88.3% of hospitalizations and 66.8% of COVID-19-related medical encounters occurred among adults at least 65 years old.

The incidence rate of COVID-19 hospitalizations per 100,000 per week was 7.38 (95% CI: 6.80, 7.99) in the unexposed cohort and 2.32 (95% CI: 2.01, 2.68) in the exposed cohort throughout the entire follow-up period. The incidence rate of any medically attended COVID-19 per 100,000 per week was 65.18 (95% CI: 63.44, 66.95) in the unexposed cohort and 41.44 (95% CI: 40.05, 42.85) in the exposed cohort.

The VE (95% CI) against COVID-19-related hospitalizations was 60.2% (53.4%–66.0%) among adults at least 18 years old (Figure 3). The VE against COVID-19-related hospitalizations was similar in the prespecified subgroups. Specifically, VE was 61.1% (54.3%–66.9%) among adults ≥50 years of age, 60.5% (53.3%–66.6%) among adults ≥65 years of age, and 58.7% (51.3%–65.0%) among adults with underlying medical conditions.

**Figure 3.**
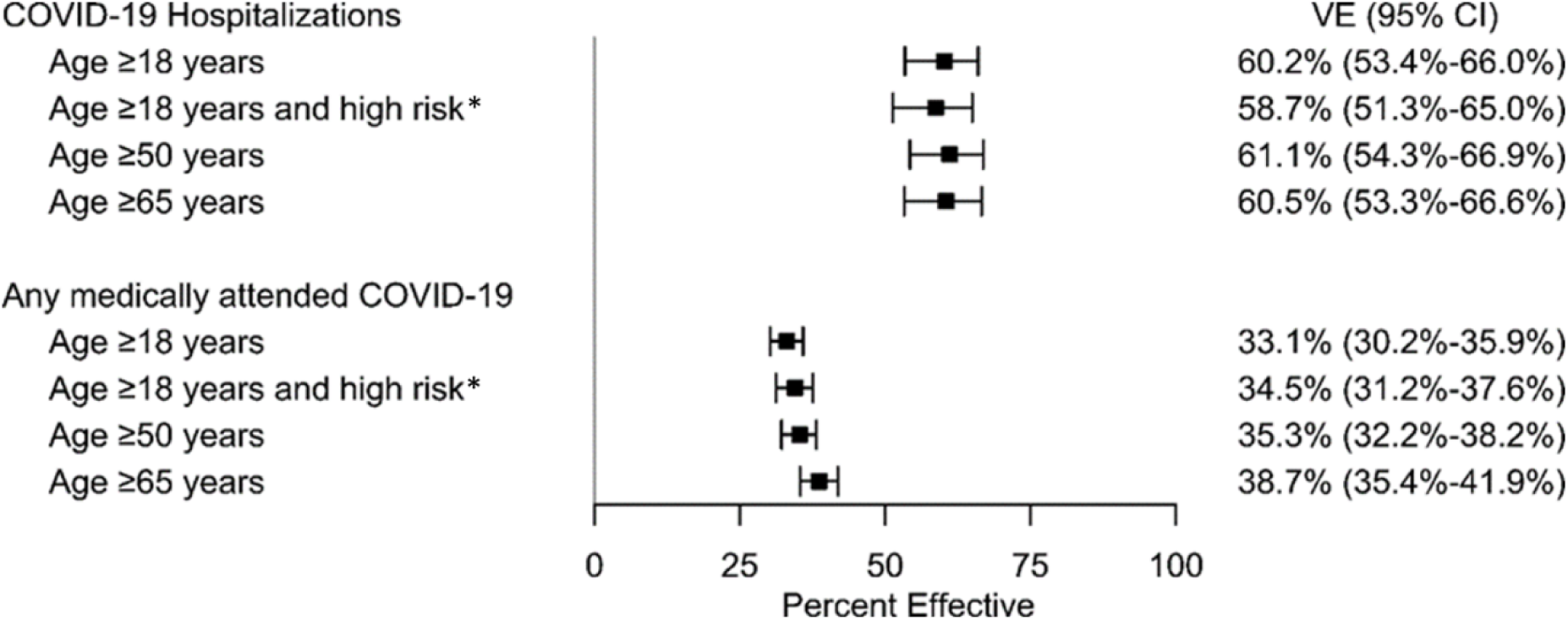
Adjusted Vaccine Effectiveness (VE) Estimates of the mRNA-1273.815 COVID-19 Vaccine. CI, confidence interval * Defined by CDC: https://www.cdc.gov/coronavirus/2019-ncov/hcp/clinical-care/underlyingconditions.html

The VE against any medically attended COVID-19 was 33.1% (30.2%–35.9%) among adults at least 18 years old. VE against medically attended COVID-19 was 35.3% (32.2%–38.2%) for adults ≥50 years old, 38.7% (35.4%–41.9%) among adults ≥65 years old, and 34.5% (31.2%–37.6%) among adults with at least one underlying medical condition.

## DISCUSSION

In this analysis of 1,718,670 US adults ≥18 years, vaccination with mRNA-1273.815, an Omicron XBB.1.5-containing COVID-19 vaccine, was associated with significant protection against COVID-19-related hospitalizations and medically attended COVID-19. Notably, the significant effectiveness of mRNA-1273.815 was observed in all subgroups, including older adults ≥ 65 and those with specific underlying medical conditions that may increase their risk of severe COVID-19 outcomes. These findings support CDC recommendations for vaccination with this 2023-2024 Omicron XBB.1.5-containing COVID-19 vaccine and underscore the benefit of receiving mRNA1273.815 for the general population and those at higher risk for COVID-19-related morbidity and mortality.

The findings of this study are consistent with other real-world effectiveness studies of Omicron XBB.1.5-containing COVID-19 vaccines conducted in the US and several European countries. Initial effectiveness studies conducted in Denmark and the Netherlands, primarily with the BNT162b2 XBB.1.5-adapted vaccine, reported effectiveness estimates of greater than 70% against COVID-19-related hospitalizations over a short follow-up period.^13,14^ Additional real-world effectiveness studies conducted in the United States using data from pharmacy, health system, and hospital-based surveillance networks estimated the effectiveness of the 2023-2024 Omicron XBB.1.5-containing mRNA COVID-19 vaccines to be ∼40% and 60% for COVID-19 outcomes, including symptomatic infection as well as illness resulting in an emergency department/urgent care visit or hospitalization.^12,15,16^ These studies had longer follow-up periods than the European studies, with a median of 30-50 days. While these studies differ in methods, population, outcomes, and duration, they are consistent with this study’s findings and demonstrate the significant benefit of vaccination with the updated 2023-2024 COVID-19 vaccines. The current study is the first to estimate effectiveness with over 60 days of follow-up post-vaccination. As the circulating variants continue to evolve, additional analyses are needed to assess the duration of the effectiveness over time.

The results of this study are also consistent with neutralizing antibody data generated among individuals who received mRNA-1273.815. A recent study in which 50 individuals received mRNA-1273.815 demonstrated a 17.5-fold increase in neutralizing antibody levels against XBB.1.5 at day 29 post-vaccination relative to pre-vaccination antibody titers.^11^ A similar increase was observed against other SARS-CoV-2 variants, including EG.5.1, XBB.1.16, BA.4/BA.5, EG.5.1, BA.2.86, and JN.1, supporting that there are significant cross-reactive immune responses from 2023-2024 Omicron XBB.1.5-containing vaccination across related SARS-CoV-2 variants. Data from a US CDC IVY study that demonstrated the effectiveness of the updated 2023-2024 vaccines reported that, in a subset of individuals in which genotyping information was available, only 16% of individuals had XBB.1.5-like spike protein mutations, with 58% having EG.5-like and 20% having HK.3-like spike mutations.^15^ The distribution of SARS-CoV-2 variants in this study was consistent with genomic surveillance conducted by the US CDC for SARS-CoV-2 during the same time period. These data suggest that while the current vaccine formulation is designed to protect against the XBB-containing SARS-CoV-2 viral variants, protection across a large number of newer viral variants has been elicited through immunologic cross-reactivity.

Four years since the start of the COVID-19 pandemic, a significant portion of the adult population in the US has developed immune responses against the SARS-CoV-2 virus, whether through previous infections, vaccinations, or a combination of both.^21^ This widespread seropositivity offers a degree of immunity within the community. However, studies have shown that the durability of this protection tends to diminish in the face of new, evolving virus variants. The current study included individuals regardless of their vaccination and infection history. The vaccinated (exposed) and the unexposed groups were matched based on prior receipt of the COVID-19 bivalent vaccine vaccination, and vaccination and infection history were adjusted for in the regression models. Thus, the result of this study provides information about the incremental protection provided by the updated vaccination in a real-world setting regardless of exposure and/or vaccination history.

The majority of the US adult population has completed their primary series vaccination. However, as of December 2023, less than 1 in 5 adults and approximately one-third of adults 65 and older had received a 2023-2024 updated COVID-19 vaccine.^8^ Studies that show the incremental protection provided by updated vaccines, especially against severe disease and among those who were previously vaccinated or infected, are essential and should be routinely communicated to increase vaccine confidence among HCPs and the general population.

### Limitations and Strengths

This study has both limitations and strengths. In terms of limitations, the study utilized EHR and claims datasets with a large sample size of insured US patients; thus, the generalizability of this study is restricted to insured US patients. In addition, inherent to most non-interventional studies utilizing real-world data, selection bias may be present due to the non-randomized nature of data collection.

Differences in patient characteristics between the two groups related to study outcomes can bias the results. To mitigate this, direct 1:1 matching and propensity score weighting were conducted using a broad range of confounders that have been shown to impact VE. After IPTW, homogenous exposed and unexposed patient groups were achieved on key measured characteristics. However, the possibility of residual unmeasured confounding persists as there may be characteristics not accounted for with the chosen study covariates, and some included covariates, such as time since last COVID-19 infection, might not be well captured in the data source if patients used an at-home test and never sought medical attention for their infection.

In addition, this study leveraged both open and closed claims for outcome ascertainment, including only patients documented in the system with continuous enrolment. While the inclusion of open claims might raise concerns regarding the incompleteness of data captured, prior studies using the same integrated dataset have shown that estimated differences of VE in open vs. closed claims were limited.^19^ Also, outcomes identification in the dataset may be limited in terms of specificity (e.g., hospitalization “due to” vs. “with” COVID-19). The selected outcome was based on a validation study of several US claims data sources in 2020, estimating the positive predictive value (PPV) of the U07.1 diagnostic code alone and in any position to range from 94.1%-81.2% for identifying COVID-19 hospitalizations.^23^ This approach aligns with prior publications of COVID-19 VE.^19,24^ The robustness of the dataset and completeness of results are also supported by the hospitalization incidence rates in this study (unexposed group), which were consistent with those reported by the CDC.^3^

Lastly, while SARS-CoV-2 still does not have a clear seasonality, this analysis included only part of the winter season, which is usually characterized by an increase in incidence rates.^22^ Partial season analyses, similar to the study presented here, aim to inform vaccine recommendations in a timely manner. Notably, this study provides the longest follow-up duration among 2023-24 COVID-19 vaccine effectiveness studies, with other published studies featuring shorter median follow-ups. Further analyses are needed to determine the durability of protection.

The strengths of this study include the use of a large, geographically distributed study population and the use of both EHR and claims data to increase the likelihood of capture of events. Furthermore, this study used both direct matching in initial patient selection and IPTW to reduce confounding between cohorts. Finally, this data source has been used extensively in vaccine effectiveness research and has been well-characterized for this application.^15^

## CONCLUSION

In this early real-world effectiveness assessment, the mRNA-1273.815 vaccine (2023-2024 Omicron XBB.1.5-containing mRNA COVID-19 vaccine) provided significant protection against COVID-19-related hospitalizations and medically-attended COVID-19. These findings are consistent with effectiveness estimates of 2023-2024 updated COVID-19 vaccines overall, demonstrating the incremental value over earlier vaccinations and the continued role of updated COVID-19 vaccinations in protection against COVID-19-related outcomes.

## Acknowledgments

Christopher Adams, an employee of Veradigm, provided programming support and quality assurance for this analysis.

## Data Availability Statement

The data that support the findings of this study were used under license from Veradigm and Komodo Health. Due to data use agreements and its proprietary nature, restrictions apply regarding the availability of the data. Further information is available from the corresponding author.

## Author Contributions

Conceptualization, J.A.M., H.K., A.B.A., D.M., M.A.M., T.L., and V.H.N.; methodology, all authors; software, N.Z.; validation, A.B., J.P.W.-J., N.Z. and I.H.W.; formal analysis, A.B., J.P.W.-J., N.Z. and I.H.W.; investigation, A.B., J.P.W.-J., N.Z., and I.H.W.; resources, M.B.; data curation, N.Z.; writing—original draft preparation, J.P.W.-J., J.A.M., and H.K.; writing—review and editing, all authors; visualization, J.P.W.-J., J.A.M. and H.K.; supervision, A.B., J.A.M., D.M., H.K. and M.B.; project administration, A.B. and M.B.; funding acquisition, J.A.M., D.M., H.K. and M.B. All authors have read and agreed to the published version of the manuscript.

## Ethics approval and consent to participate

The linked dataset only contains de-identified data as per the de-identification standard defined in Section §164.514(a) of the Health Insurance Portability and Accountability Act of 1996 (HIPAA) Privacy Rule. The process by which the data is de-identified is attested to through a formal determination by a qualified expert as defined in Section §164.514(b)(1) of the HIPAA Privacy Rule. Because this study used only de-identified patient records, it is therefore no longer subject to the HIPAA Privacy Rule and is therefore exempt from Institutional Review Board approval and for obtaining informed consent according to US law. This study was conducted in compliance with the Declaration of Helsinki and used only de-identified data.

## Supplementary Tables

**Supplementary Table 1.** List of codes used to identify 2023-2024 Omicron XBB.1.5-containing COVID-19 vaccines from the Veradigm EHR and linked claims datasets.

| COVID-19 vaccine type | CPT | CVX | NDC |
| --- | --- | --- | --- |
| mRNA-1273.815 | 91321, 91322 | 311, 312 | 80777-0102-01, 80777-0102-04, 80777-0102-93, 80777-0102-95, 80777-0102-96, 80777-0287-07, 80777-0287-92 |
| BNT162b2 XBB.1.5-adapted vaccine | 91318, 91319, 91320 | 308, 309, 310 | 00069-2362-01, 00069-2362-10, 00069-2392-01, 00069-2392-10, 59267-4315-01, 59267-4315-02, 59267-4331-01, 59267-4331-02 |
| 2023-2024 Novavax updated vaccine |  |  | 80631-0105-01, 80631-0105-02 |

CPT, Current Procedural Terminology; CVX, vaccine administered codes; NDC, National Drug Codes

**Supplementary Table 2.** List of CVX, CPT, and NDC codes used to identify COVID-19 vaccines from the Veradigm EHR and linked claims datasets.

| Manufacturer | CPT | CVX | NDC | ICD-10-PCS |
| --- | --- | --- | --- | --- |
| AstraZeneca | 91302, 0021A, 0022A | 210 | 00310-1222-10, 00310-1222-15 |  |
| Janssen | 91303, 0031A, 0034A | 212 | 59676-0580-05, 59676-0580-15 |  |
| Moderna | 91301, 91306, 91309, 91311, 91313, 91314, 91316, 91321, 91322, 0011A, 0012A, 0013A, 0064A, 0091A, 0092A, 0093A, 0094A, 0111A, 0112A, 0113A, 0134A, 0141A, 0142A, 0144A, 0164A | 207, 221, 227, 228, 229, 230, 311, 312, 519 | 61434-0043-02, 80777-0100-11, 80777-0100-15, 80777-0100-98, 80777-0100-99, 80777-0102-01, 80777-0102-04, 80777-0102-93, 80777-0102-95, 80777-0102-96, 80777-0273-10, 80777-0273-15, 80777-0273-98, 80777-0273-99, 80777-0275-05, 80777-0275-99, 80777-0277-05, 80777-0277-99, 80777-0279-05, 80777-0279-99, 80777-0280-05, 80777-0280-99, 80777-0282-05, 80777-0282-99, 80777-0283-02, 80777-0283-99, 80777-0287-07, 80777-0287-92 |  |
| Novavax | 91304, 0041A, 0042A, 0044A | 211, 313 | 80631-0100-01, 80631-0100-10, 80631-0102-01, 80631-0105-01, 80631-0105-02, 80631-1000-01 |  |
| Pfizer | 91317, 91318, 91319, 91320, 0121A, 0151A, 0171A, 0172A, 0173A, 91300, 91305, 91307, 91308, 91312, 91315, 0001A, 0002A, 0003A, 0004A, 0051A, 0052A, 0053A, 0054A, 0071A, 0072A, 0073A, 0074A, 0081A, 0082A, 0083A, 0124A, 0154A | 219, 300, 301, 302, 308, 309, 310, 520, 208, 217, 218 | 00069-1000-01, 00069-1000-02, 00069-1000-03, 00069-2025-01, 00069-2025-10, 00069-2025-25, 00069-2362-01, 00069-2362-10, 00069-2392-01, 00069-2392-10, 59267-0078-01, 59267-0078-02, 59267-0078-04, 59267-0304-01, 59267-0304-02, 59267-0565-01, 59267-0565-02, 59267-0609-01, 59267-0609-02, 59267-1000-01, 59267-1000-02, 59267-1000-03, 59267-1025-01, 59267-1025-02, 59267-1025-03, 59267-1025-04, 59267-1055-01, 59267-1055-02, 59267-1055-04, 59267-1404-01, 59267-1404-02, 59267-4315-01, 59267-4315-02, 59267-4331-01, 59267-4331-02 |  |
| Sanofi | 91310, 0104A | 225, 226 | 49281-0618-20, 49281-0618-78 |  |
| Not Specified | M0201 | 213 |  | XW013S6, XW013T6, XW013U6, XW023S6, XW023T6, XW023U6 |

CPT, Current Procedural Terminology; CVX, vaccine administered codes; ICD-10-PCS, International Classification of Disease, 10<sup>th</sup> edition, Procedure Coding System; NDC, National Drug Codes

**Supplementary Table 3.**
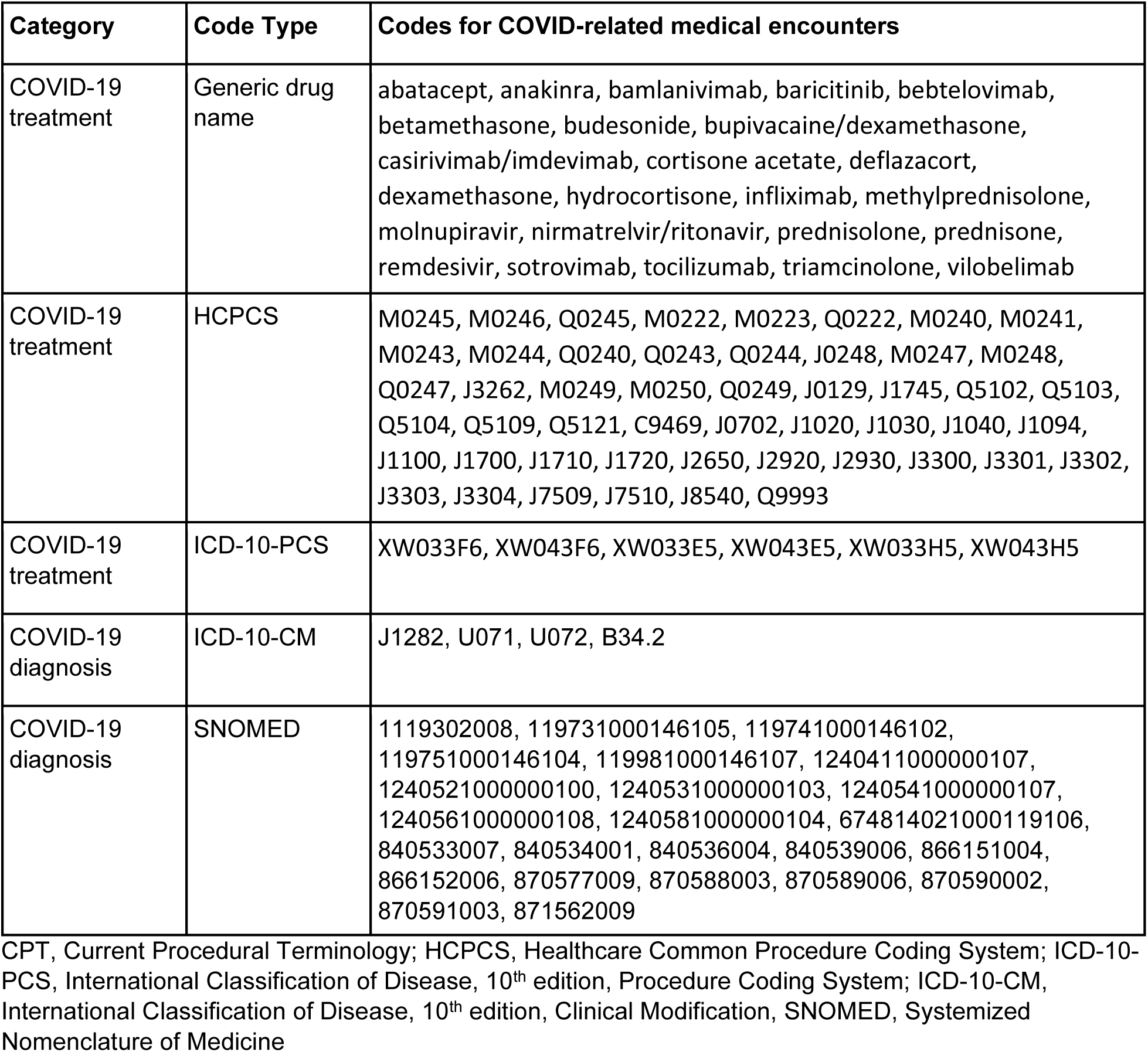
Codes used to identify COVID-19 diagnosis or treatment.

| Category | Code Type | Codes for COVID-related medical encounters |
| --- | --- | --- |
| COVID-19 treatment | Generic drug name | abatacept, anakinra, bamlanivimab, baricitinib, bebtelovimab, betamethasone, budesonide, bupivacaine/dexamethasone, casirivimab/imdevimab, cortisone acetate, deflazacort, dexamethasone, hydrocortisone, infliximab, methylprednisolone, molnupiravir, nirmatrelvir/ritonavir, prednisolone, prednisone, remdesivir, sotrovimab, tocilizumab, triamcinolone, vilobelimab |
| COVID-19 treatment | HCPCS | M0245, M0246, Q0245, M0222, M0223, Q0222, M0240, M0241, M0243, M0244, Q0240, Q0243, Q0244, J0248, M0247, M0248, Q0247, J3262, M0249, M0250, Q0249, J0129, J1745, Q5102, Q5103, Q5104, Q5109, Q5121, C9469, J0702, J1020, J1030, J1040, J1094, J1100, J1700, J1710, J1720, J2650, J2920, J2930, J3300, J3301, J3302, J3303, J3304, J7509, J7510, J8540, Q9993 |
| COVID-19 treatment | ICD-10-PCS | XW033F6, XW043F6, XW033E5, XW043E5, XW033H5, XW043H5 |
| COVID-19 diagnosis | ICD-10-CM | J1282, U071, U072, B34.2 |
| COVID-19 diagnosis | SNOMED | 1119302008, 119731000146105, 119741000146102, 119751000146104, 119981000146107, 1240411000000107, 1240521000000100, 1240531000000103, 1240541000000107, 1240561000000108, 1240581000000104, 674814021000119106, 840533007, 840534001, 840536004, 840539006, 866151004, 866152006, 870577009, 870588003, 870589006, 870590002, 870591003, 871562009 |

CPT, Current Procedural Terminology; HCPCS, Healthcare Common Procedure Coding System; ICD-10-PCS, International Classification of Disease, 10<sup>th</sup> edition, Procedure Coding System; ICD-10-CM, International Classification of Disease, 10<sup>th</sup> edition, Clinical Modification, SNOMED, Systemized Nomenclature of Medicine

**Supplementary Table 4.**
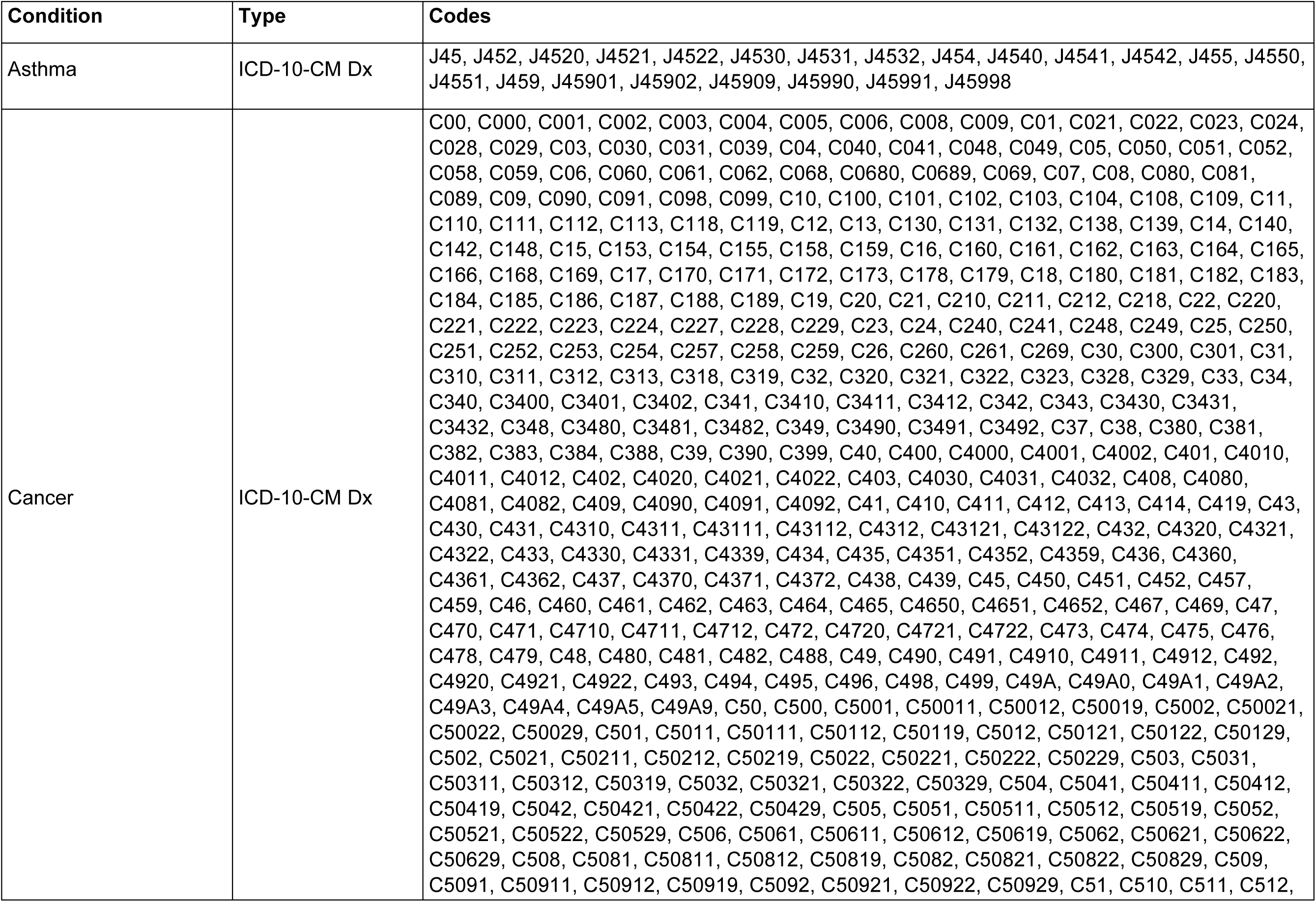
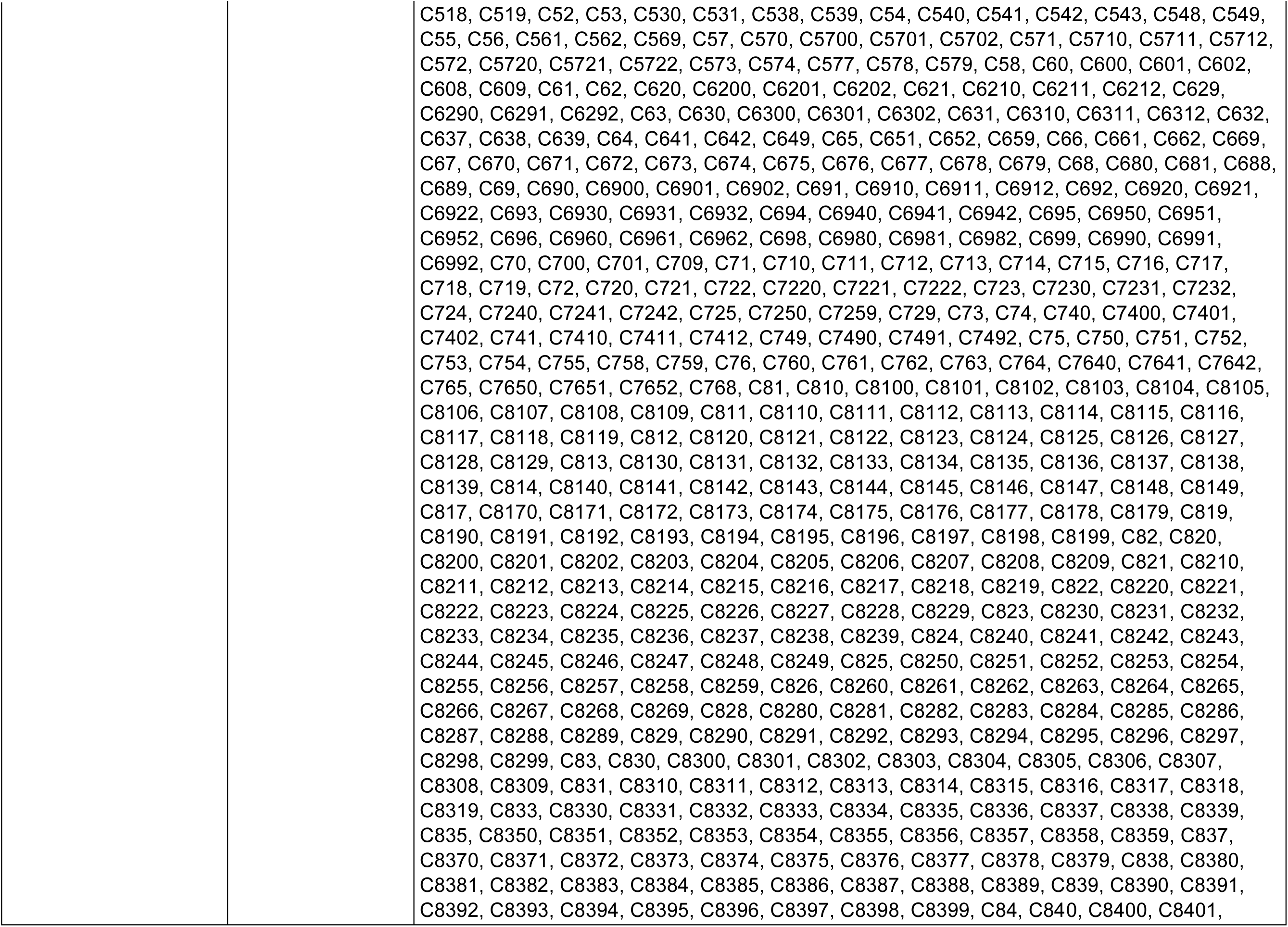
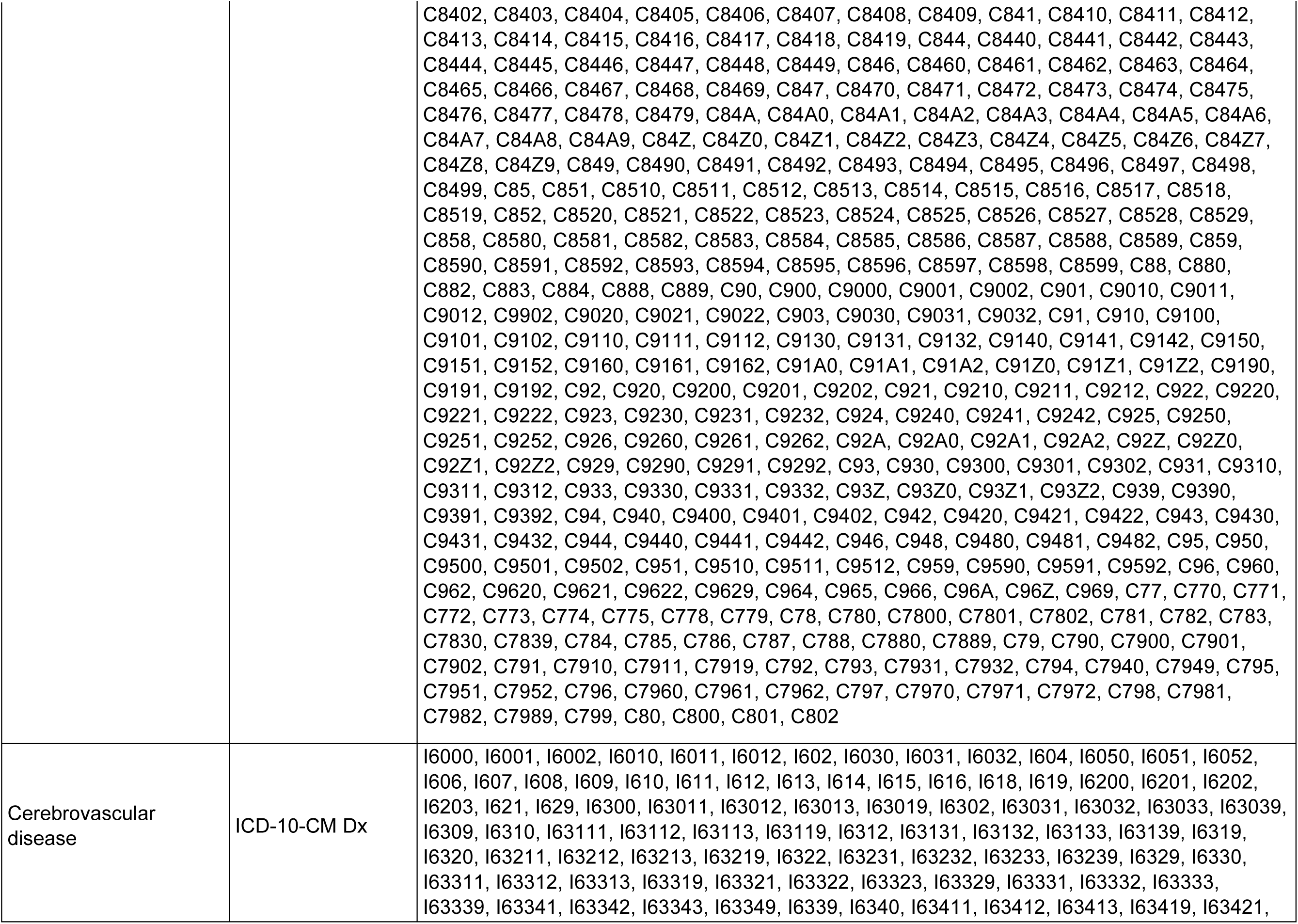
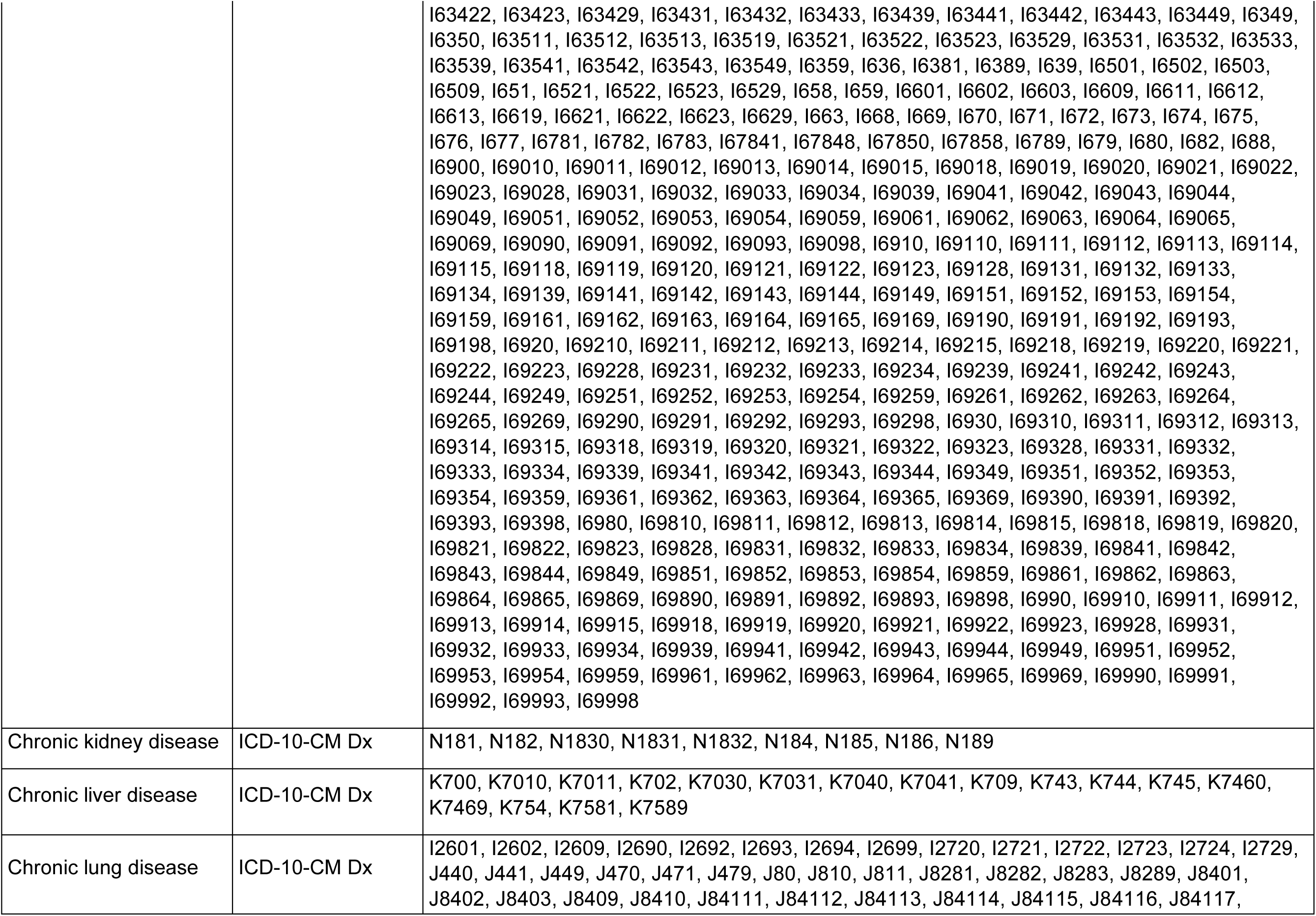
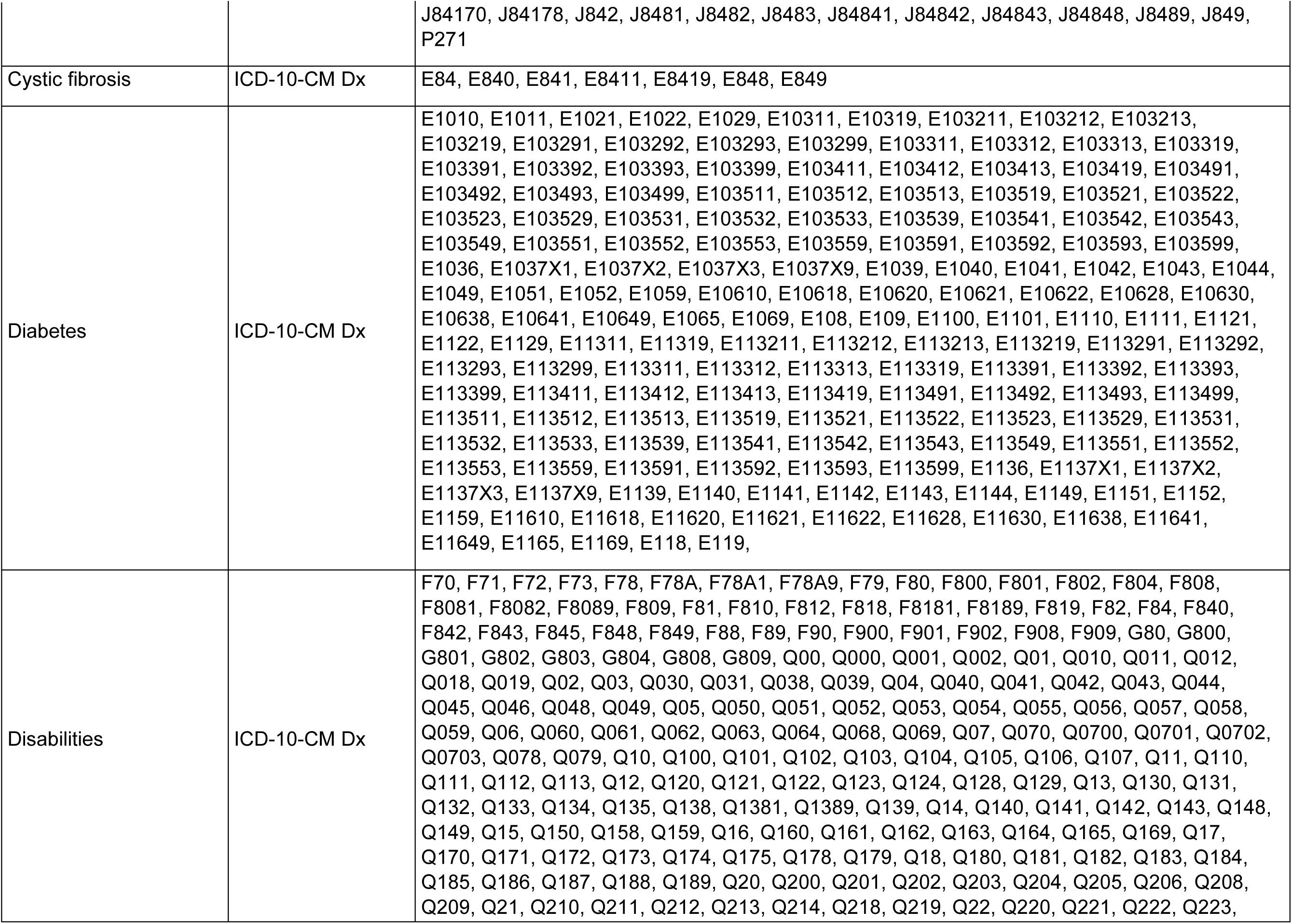
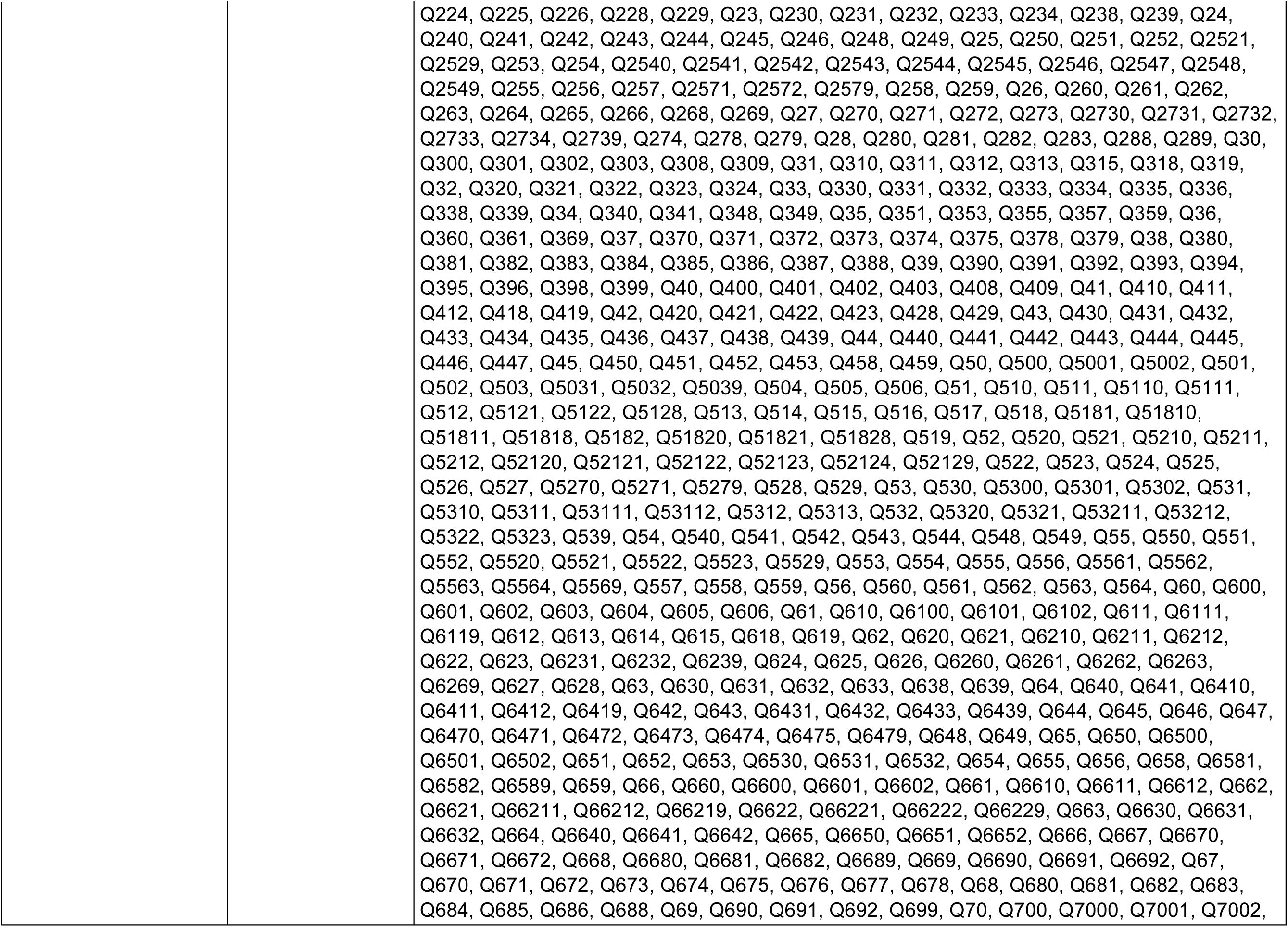
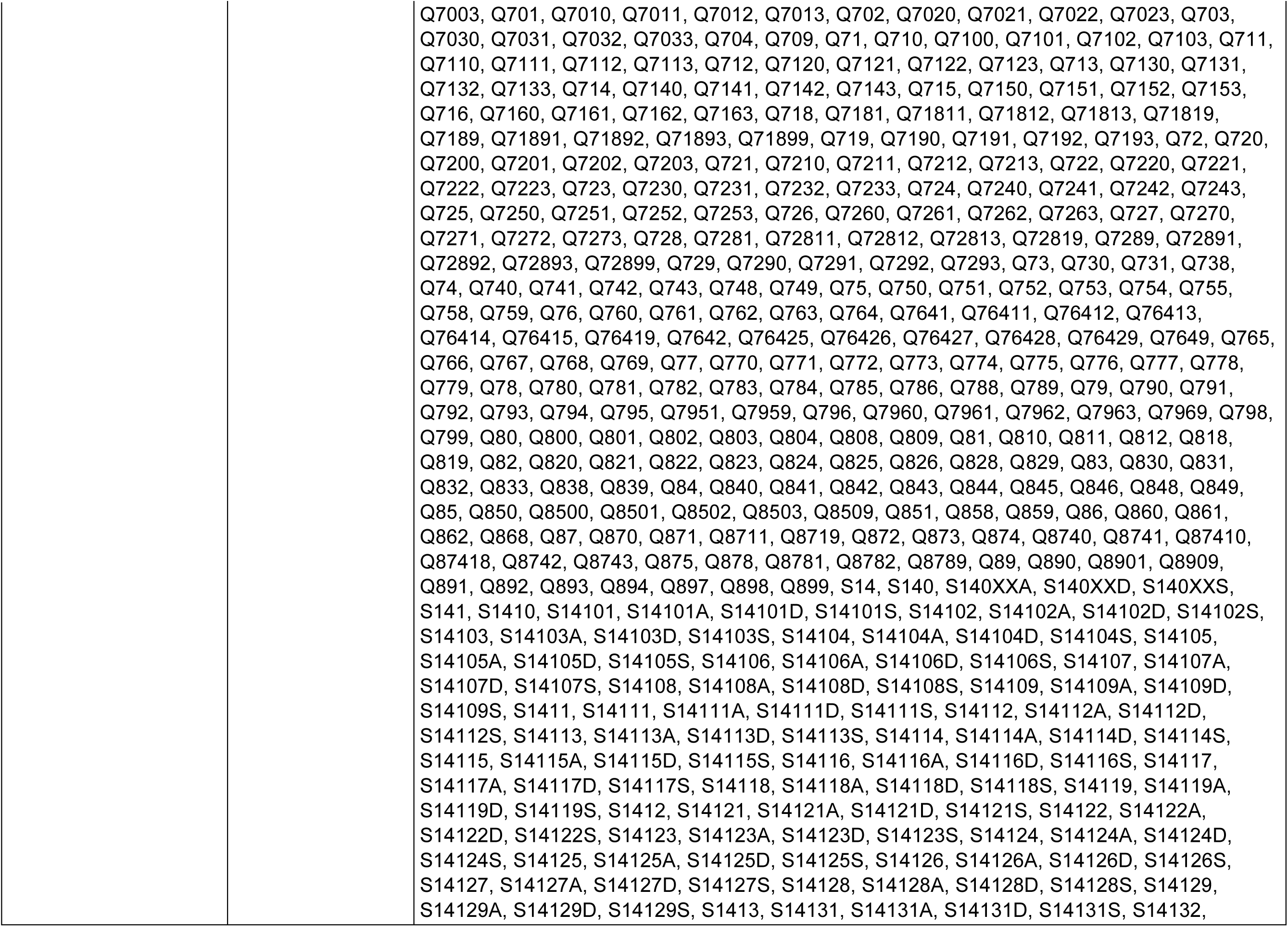
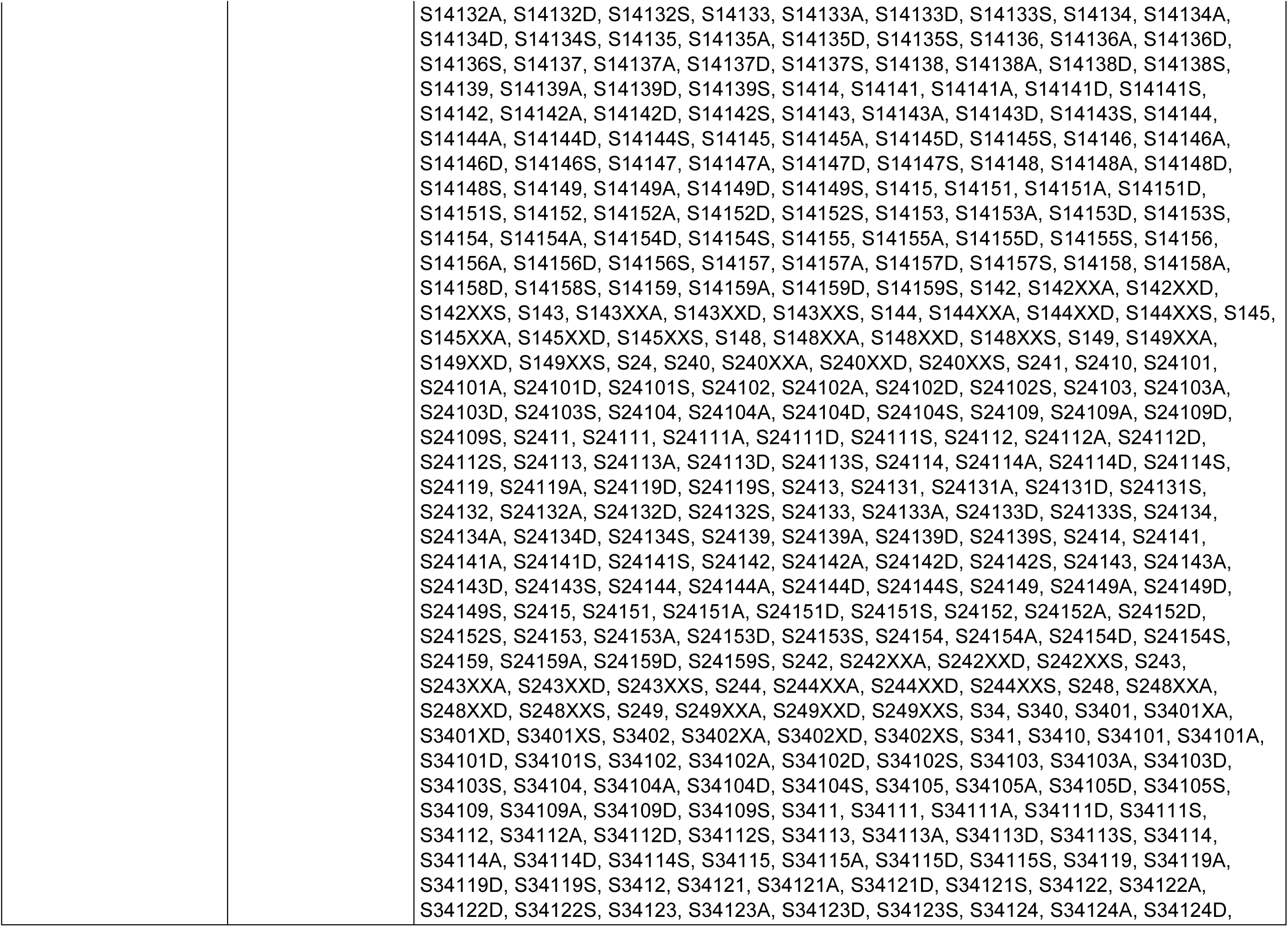
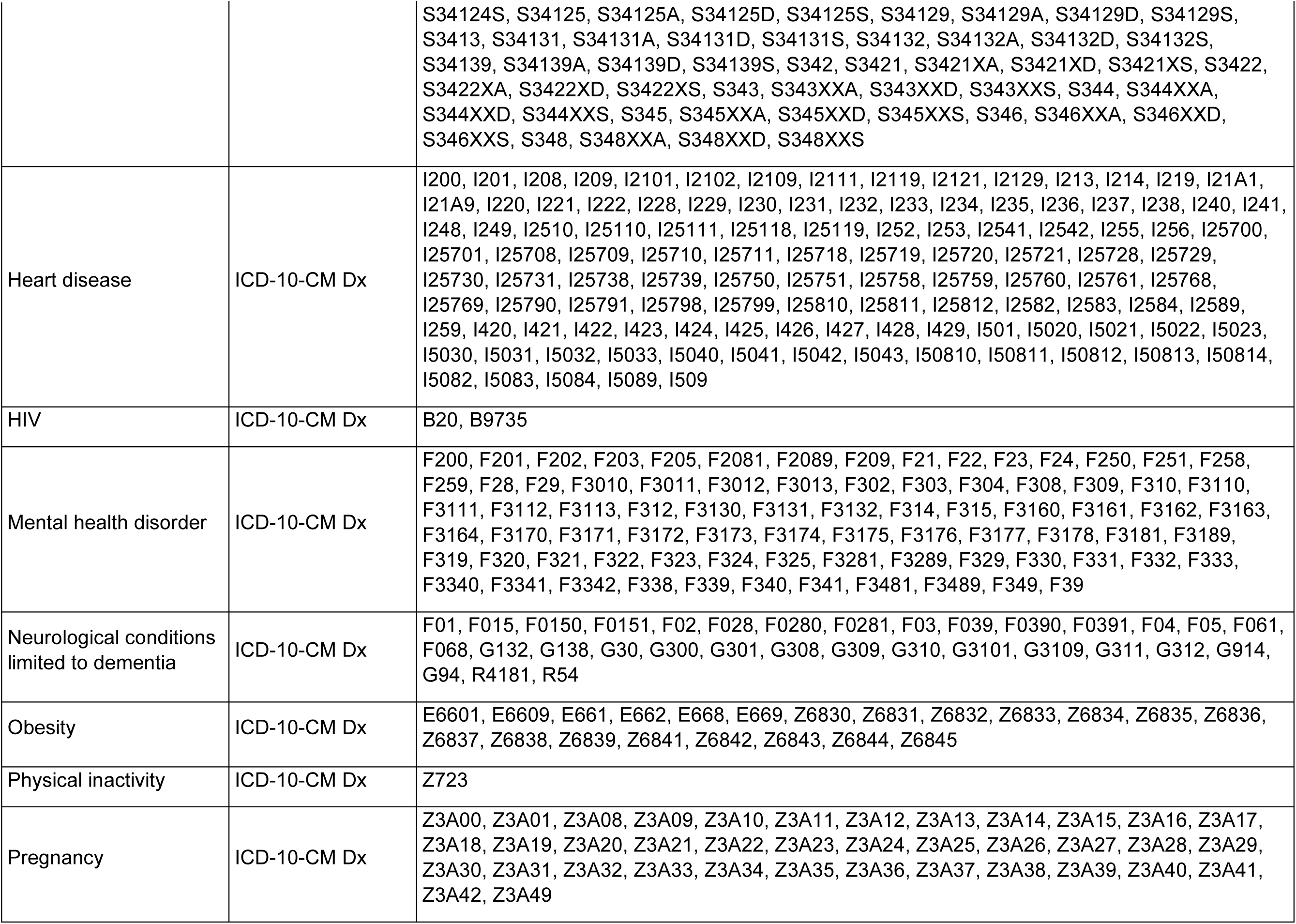
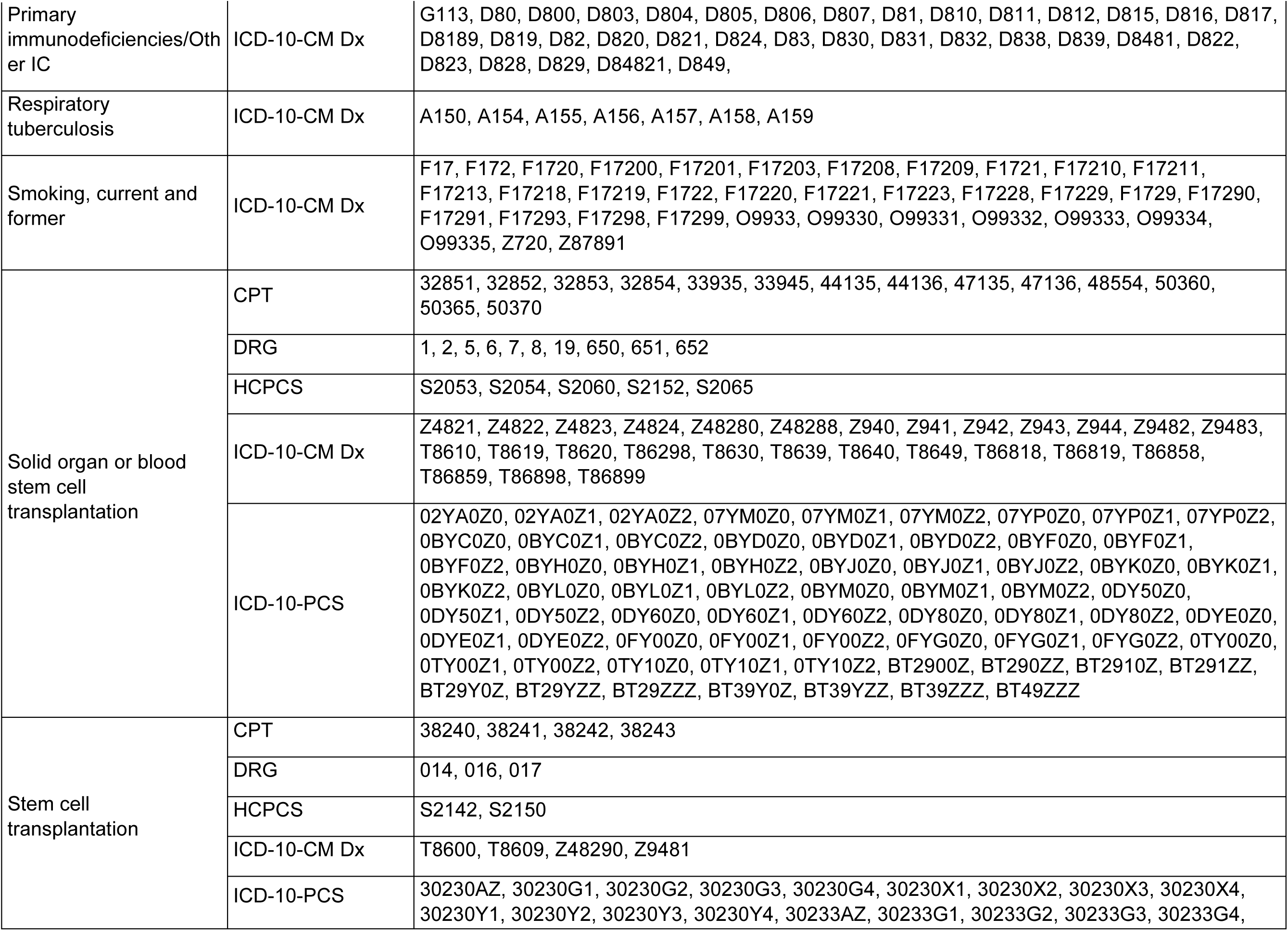
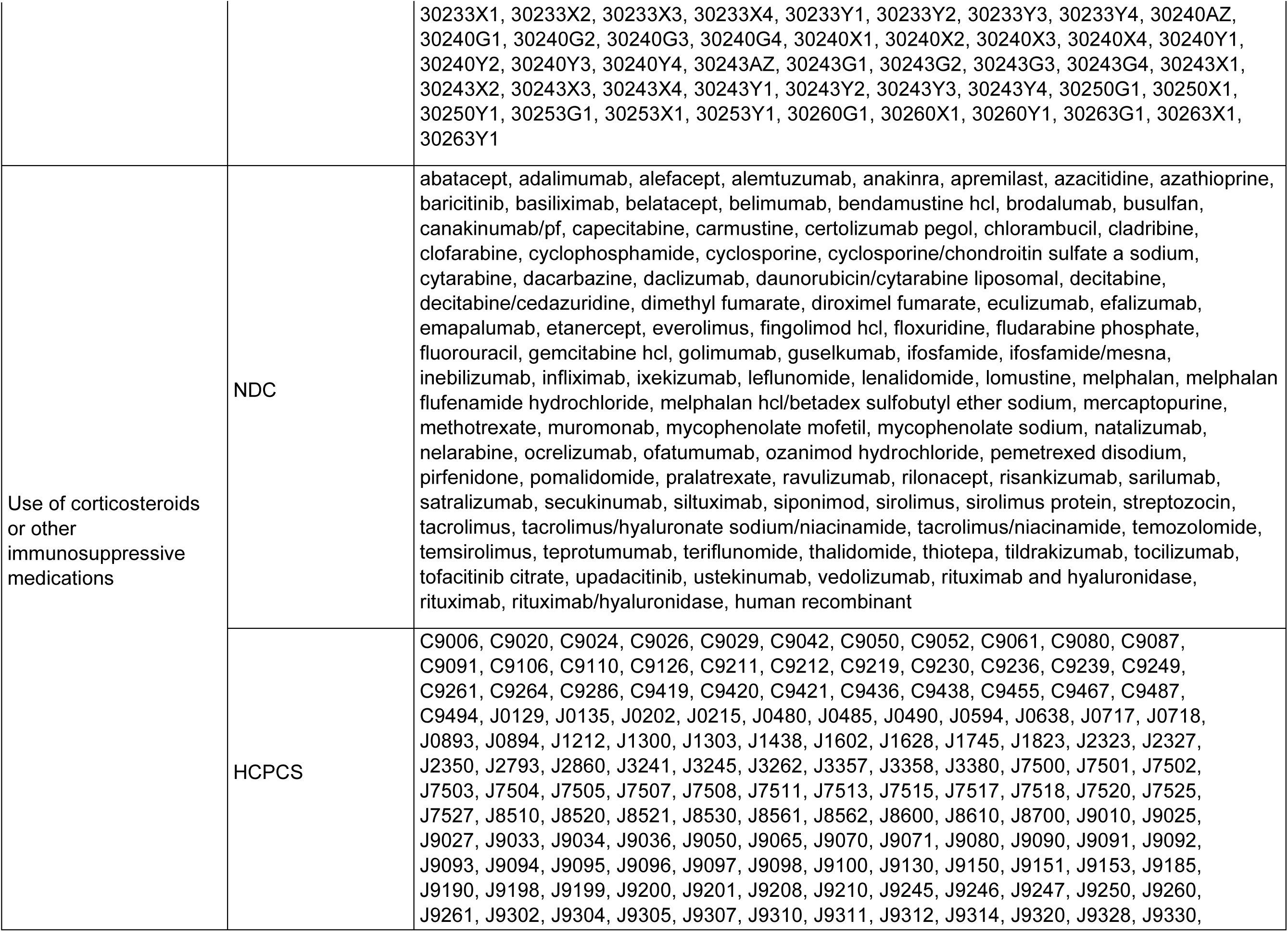
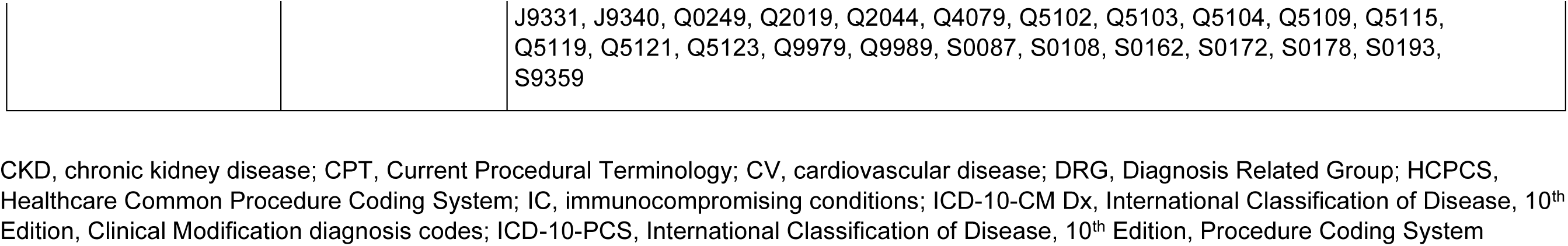
Diagnostic codes used for identifying patients with underlying medical conditions.

